# Vibratory stimulation and not action observation or mirror therapy reduces paralyzed hand spasticity in chronic stroke survivors

**DOI:** 10.1101/2024.12.03.24318436

**Authors:** Hideki Nakano, Kotaro Nakagawa, Tatsuya Ikuno, Munenori Nagashima, Eiichi Naito

**Affiliations:** Center for Information and Neural Networks (CiNet), Advanced ICT Research Institute, National Institute of Information and Communications Technology (NICT), Suita, Japan; Graduate School of Health Sciences, Kyoto Tachibana University, Kyoto, Japan; Department of Physical Therapy, Faculty of Health Sciences, Kyoto Tachibana University, Kyoto, Japan; Nagashima Neurosurgery Rehabilitation Clinic, Osaka, Japan; Ugokinokotsu Institute Rehabilitation Center, Nishinomiya, Japan; Graduate School of Frontier Biosciences, Osaka University, Suita, Japan

**Author notes:** ***Corresponding author:*** Eiichi Naito, PhD.

**Keywords:** stroke, hand spasticity, vibratory stimulation, action observation, mirror therapy

## Abstract

**Objective:** Post-stroke spasticity prevalence increases with time from stroke onset, and negatively impacts the patient’s quality of life. Vibratory stimulation (VS) to the spastic muscles can reduce spasticity, whereas action observation (AO) and mirror therapy (MT) are reported to be less effective for spasticity reduction. However, these effects have not been compared within the same individuals. Therefore, this study compared the spasticity-reducing effects of VS, AO, and MT.

**Methods:** Independent groups of 13 patients with chronic stroke participated in the study. In Experiment 1, the participants underwent VS, AO, and combined VS+AO on separate days. In Experiment 2, different participants received VS, MT, and VS+MT. Spasticity was assessed using the Modified Ashworth Scale (MAS) before and after the interventions.

**Results:** In Experiment 1, VS significantly reduced MAS scores but not AO, and AO did not enhance the VS effect in combined therapy. In Experiment 2, VS significantly reduced the MAS scores, but not MT, and MT did not enhance the VS effect in combined therapy.

**Conclusion:** Spastic muscle VS reduced spasticity in chronically paralyzed hands; however, AO and MT did not enhance the VS effect. VS alone may be a more effective treatment for spasticity in patients with chronic stroke.

## Introduction

Post-stroke spasticity is a movement disorder characterized by a velocity-dependent increase in the tonic stretch reflex with increased tendon reflexes (1). Spasticity is more frequently observed in the upper limbs than the lower limbs (2), and hinders motor function recovery (3); therefore, effective interventions are needed to reduce spasticity in neurorehabilitation.

The post-stroke spasticity prevalence has been reported to be 4–27% at 14 weeks after stroke, 1–26.7% at 13 months after stroke, and 17–42.6% at > 3 months after stroke (4). Thus, the post-stroke spasticity prevalence increases with time from stroke onset (5). Persistent spasticity in patients with chronic stroke has a significant negative impact on their activities of daily living and health-related quality of life (6). Therefore, understanding the effects of reducing spasticity, in patients with chronic stroke is important from the perspective of spasticity management and treatment.

Vibratory stimulation (VS), a nonpharmacological treatment for post-stroke spasticity, is recommended in guidelines as a spasticity reducing method in patients with stroke (7,8). Furthermore, a recent systematic review and meta-analysis revealed that VS is effective in reducing spasticity in patients with stroke (9). Previous studies have reported that short-term VS applied directly to the upper limb spastic muscles of patients with stroke can reduce spasticity (10–12). Noma et al. administered a single 5-min VS on the spastic flexor muscles of the fingers, wrist, and elbow in patients with stroke and revealed that muscle spasticity was significantly reduced immediately after the intervention (10,11). Takeuchi et al. also reported that 5-min VS administration to the spastic flexor muscles of the fingers and wrists in patients with stroke significantly reduced their spasticity. Furthermore, VS on the tendon reduced spasticity more than that on the muscle belly (12). However, these effects have been confirmed during the subacute phase (< 6 months after stroke onset) (13) and have never been confirmed during the chronic phase (> 6 months).

Action observation (AO) and mirror therapy (MT) have been widely used to restore upper limb motor function in patients with stroke (14,15). The effects of AO and MT on spasticity in paralyzed hands of patients with stroke have been examined in several randomized controlled trials (RCTs). As for AO, Mancuso M et al. (16) investigated its effects on patients with acute stroke for approximately 15 days. Furthermore, Kim CH et al. (17) and Franceschini M et al. (18) conducted similar studies for 4 weeks in patients with subacute stroke. As for MT, Samuelkamaleshkumar S et al. (19) characterized the effect of MT for 3 weeks in patients with acute stroke, Yavuzer G et al. (20) for 4 weeks, and Mirela Cristina L et al. (21) for 6 weeks in patients with subacute stroke. Furthermore, Michielsen ME et al. (22) assessed the MT effects for 6 weeks in patients with chronic stroke. All these RCTs reported that AO and MT improved upper limb motor function or activities of daily living; however, none of them reported significant spasticity reduction.

Thus, VS is effective for post-stroke spasticity; while, the effects of AO and MT are limited. However, the effects of VS, AO, and MT have not been compared in the same individuals. Therefore, this study compared the spasticity-reducing effects of VS, AO, and MT in patients with stroke to assess the effectiveness of VS in the chronic phase.

## Material and methods

### Participants

Overall, 13 patients with chronic stroke (male = 7, female = 6, mean age 60.38 ± 7.81 years) participated in Experiment 1, and other 13 patients with chronic stroke (male = 9, female = 4, mean age 55.62 ± 7.68 years) participated in Experiment 2. Inclusion criteria were patients with first stroke > 6 months after onset, and patients with spasticity of the paralyzed hand and a Modified Ashworth Scale (MAS) score of ≥ 1 (23,24). The exclusion criteria were orthopedic, psychiatric, and cardiovascular diseases that may affect the results of this study, and severe sensory impairment, pain, numbness, higher brain dysfunction, and cognitive dysfunction. The participant characteristics are presented in Table 1.

**Table 1.** Participant characteristics.

|  |  | Experiment 1<br>(n = 13) | Experiment 2<br>(n = 13) |
| --- | --- | --- | --- |
| Male / Female (n) |  | 7 / 6 | 9 / 4 |
| Age (year) |  | 60.38 ± 7.81 | 55.62 ± 7.68 |
| Time since onset (month) |  | 49.31 ± 27.81 | 26.69 ± 17.87 |
| Stroke type | Hemorrhagic / Ischemic (n) | 8 / 5 | 10 / 3 |
| Paralyzed side | Right / Left (n) | 5 / 8 | 8 / 5 |
| FMA hand (score) |  | 6.08 ± 5.04 | 7.08 ± 3.86 |
| SIAS UE touch (score) |  | 1.85 ± 0.99 | 2.00 ± 0.71 |
| SIAS UE position (score) |  | 1.92 ± 1.26 | 2.08 ± 1.19 |
| MAS (score) |  | 2.46 ± 1.33 | 2.08 ± 1.04 |

Mean ± SD; FMA, Fugl-Meyer Assessment; SIAS, Stroke Impairment Assessment Set; UE, Upper Extremity; MAS, Modified Ashworth Scale.

This study was conducted in accordance with the Declaration of Helsinki and informed consent was obtained from all participants. This study was approved by the Kyoto Tachibana University Research Ethics Committee (approval number 21-47). This study was registered in the UMIN Clinical Trials Registry (UMIN000056192).

### Experiment 1

In Experiment 1, the participants were randomly assigned to receive VS, AO, or and VS+AO on different days. A washout period of approximately one week was maintained between each intervention (VS, AO, or VS+AO).

#### VS: Vibratory stimulation (Figure 1A)

The participants sat on a chair with a backrest and placed both upper limbs on a desk in a relaxed posture. The participants were then asked to grasp a vibration stimulator (PLANSTAFF. Co., Ltd., Japan) (25) using the paralyzed hand. The vibration stimulator was adjusted to contact each finger and palm of the paralyzed hand. Participants were verbally instructed to relax their paralyzed hand. High-frequency (80-120 Hz) and low-amplitude VS has been reported to induce the activity of the Ia afferent nerves from the muscle spindles (26–28). Therefore, the VS in this study was set to a frequency of 80 Hz and an amplitude of 1.0 mm. VS was performed with the eyes closed for 5 min.

**Figure 1.**
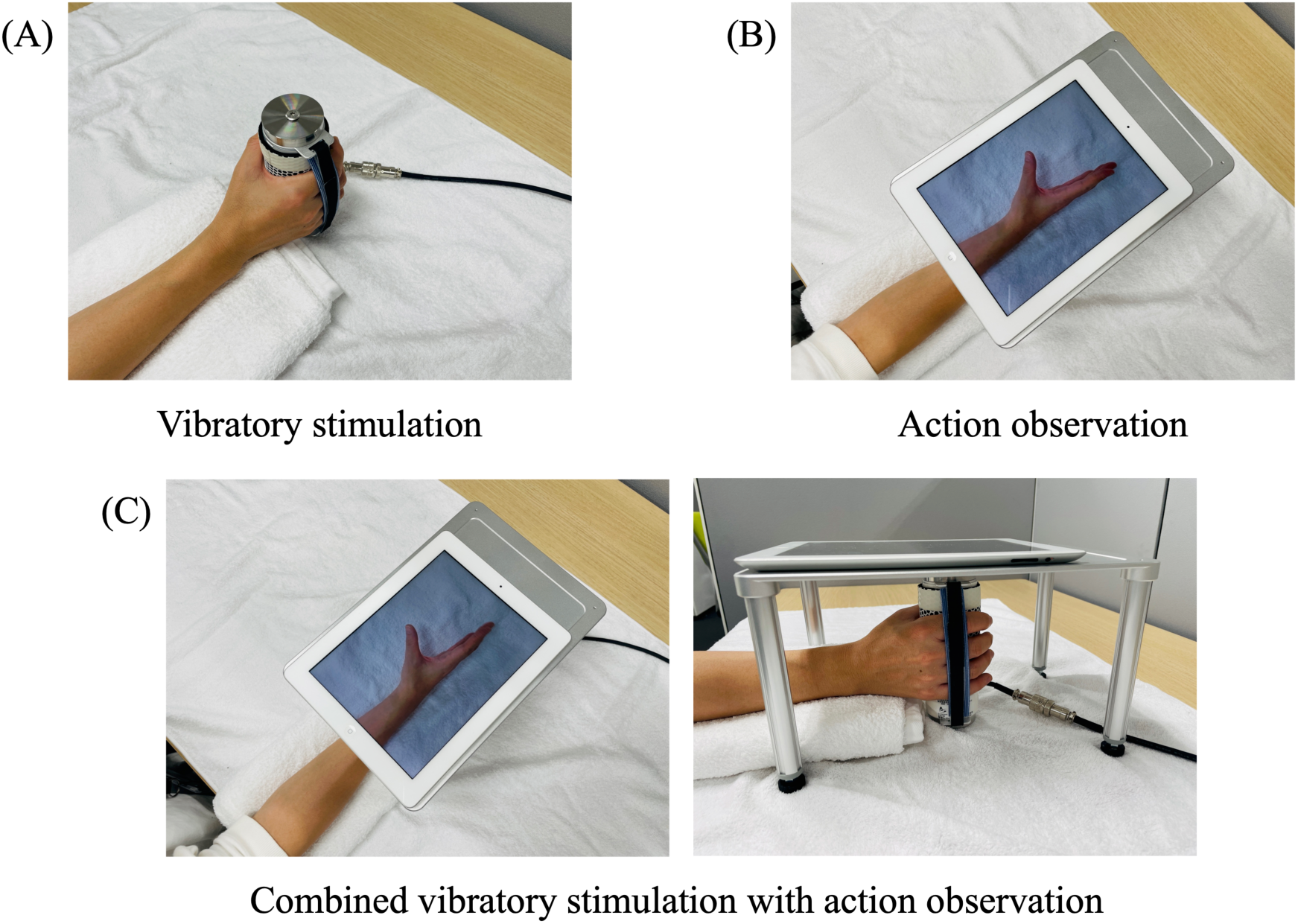
Interventions in Experiment 1. (A) Vibratory stimulation. Vibratory stimulation was provided to the fingers and the palm of the paralyzed hand using a vibration stimulator. (B) Action observation. A tablet was placed over the paralyzed hand, and the flexion and extension movements of the participant’s healthy fingers were displayed on the tablet. (C) Combined vibratory stimulation with action observation. A tablet was placed over the paralyzed hand, lightly holding a vibration stimulator.

#### AO: Action observation (Figure 1B)

The participants sat on a chair with backrest and placed both upper limbs on a desk in a relaxed posture. Before we started AO, we prepared a video. A therapist (KN) used a tablet with a camera (Apple, USA) to film the flexion and extension movements of the healthy fingers. The participants were then asked to hold the healthy hand in a neutral forearm position with all fingers in a flexed position. Next, they were asked to extend all fingers from the flexed position for 3 s, and then to flex all fingers from the extended position for 3 s. This 6-second flexion and extension movement (3-s extension phase, 3-s flexion phase) was performed thrice consecutively, while the therapist recorded the movement from above. The video footage was flipped to the left and right using an application (Flip video & Rotate video app, Sounak Sarkar). During AO, the tablet was placed over the participant’s paralyzed hand, and the flipped video was played continuously using an application (Infinite Loop Player, Airwire products). The paralyzed forearm position was adjusted to match that of the healthy forearm in the video. The participants were verbally instructed to observe the flexion and extension movements of the fingers on the tablet, as if they were moving their paralyzed hands themselves. Thus, motor imagery component was included during AO. AO was performed for 5 min.

#### VS+AO: Combined vibratory stimulation with action observation (Figure 1C)

The participants sat on a chair with a backrest and placed both upper limbs on a desk in a relaxed posture. The VS was performed described above. Simultaneously, AO was performed as described above. VS+AO was performed with the eyes open for 5 min.

### Experiment 2

In Experiment 2, the participants were randomly assigned to receive VS, MT, and VS+MT on different days. A washout period of approximately one week was maintained between each intervention (VS, MT, or VS+MT).

#### VS: Vibratory stimulation (Figure 2A)

As in the Experiment 1, VS was performed with eyes closed for 5 min.

**Figure 2.**
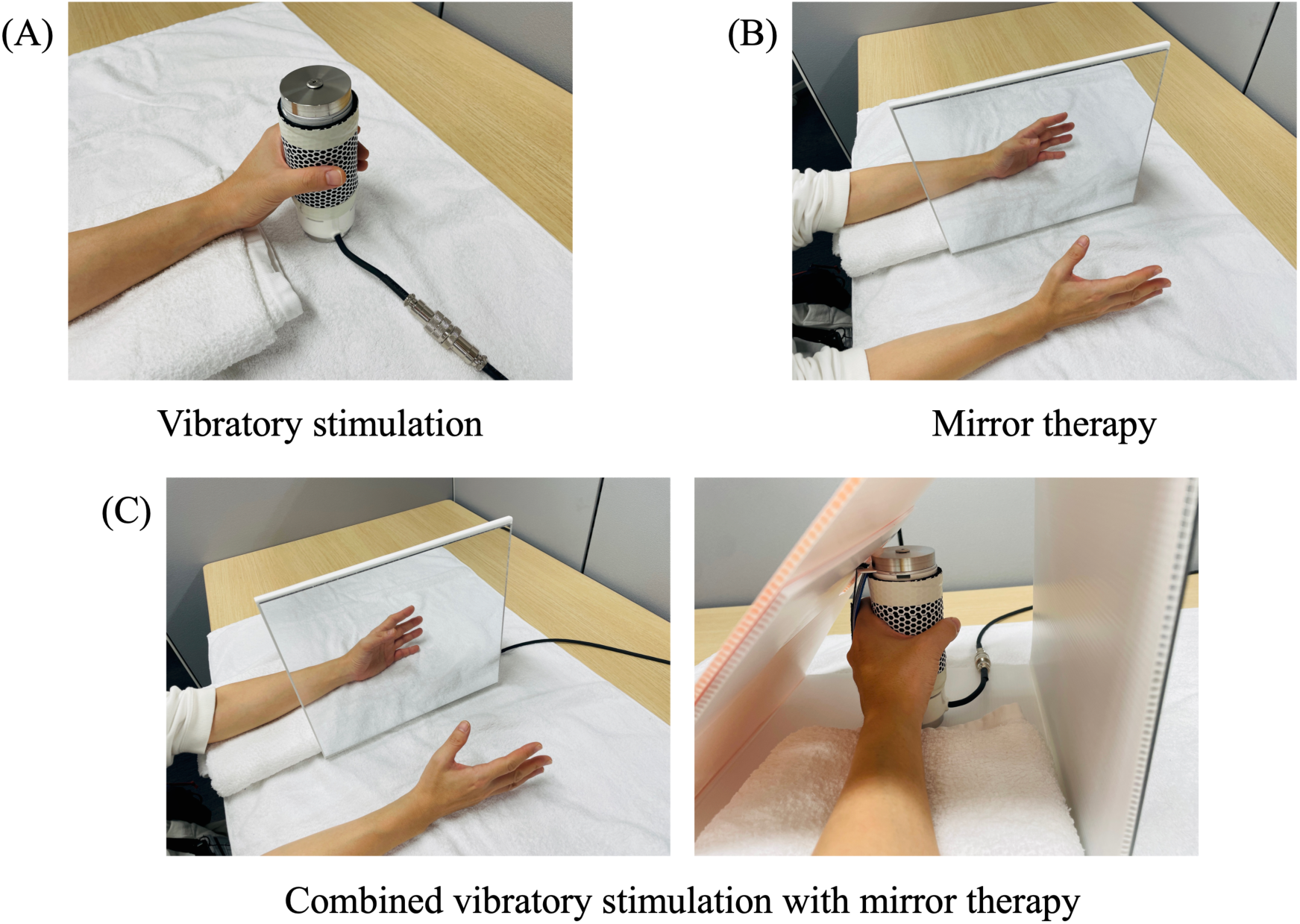
Interventions in Experiment 2. (A) Vibratory stimulation. Vibratory stimulation was provided to the fingers and the palm of the paralyzed hand using a vibration stimulator. (B) Mirror therapy. A mirror was placed in front of the paralyzed hand, and the participant slowly flexed and extended the healthy fingers. (C) Combined vibratory stimulation with mirror therapy. A mirror was placed in front of the paralyzed hand, lightly holding a vibration stimulator.

#### MT: Mirror therapy (Figure 2B)

The participants sat on a chair with a backrest and placed both upper limbs on a desk in a relaxed posture. The participants placed their paralyzed hand in a mirror box (Noigroup, Australia) with a neutral forearm position. The healthy hand was placed on the opposite side of the mirror box with a neutral forearm position and all fingers flexed. The participant was then asked to repeatedly perform flexion and extension movements of the healthy fingers (3-s extension phase, 3-s flexion phase). The participants were verbally instructed to observe the flexion and extension movements of the fingers reflected in the mirror as if they were moving their paralyzed hands themselves. Thus, motor imagery component was included during MT. The MT was performed for 5 min.

#### VS+MT: Combined vibratory stimulation with mirror therapy (Figure 2C)

The participants sat on a chair with a backrest and placed both upper limbs on a desk in a relaxed posture. The VS was performed as described above. Moreover, MT was performed as described above. VS+MT was performed with the eyes open for 5 min.

### Measures

The spasticity degree of the paralyzed finger flexor muscles was assessed before and after each intervention. MAS is used as an assessment index of spasticity (23,24). The reliability of the MAS has been established in a previous systematic review and meta-analysis (24). The MAS had a 6-point scale (0, “no increase in muscle tone”; 1, “slight increase in muscle tone, manifested by a catch and release or by minimal resistance at the end of the range of motion when the affected part(s) is moved in flexion or extension”; 1+, “slight increase in muscle tone, manifested by a catch, followed by minimal resistance throughout the remainder (less than half) of the range of motion (ROM)”; 2, “more marked increase in muscle tone through most of the ROM, but affected part(s) easily moved”; 3, “considerable increase in muscle tone, passive movement difficult”; and 4, “affected part(s) rigid in flexion or extension”.) (23) In the data analysis, the MAS scores (0, 1, 1+, 2, 3, 4) were converted to discrete categorical scores (0, 1, 2, 3, 4, 5) (10–12). All assessments were performed by an occupational therapist with 20 years of clinical experience (KN) to avoid interrater variability.

### Statistical analysis

A non-parametric two-way ANOVA with an aligned rank transform (ART ANOVA) was used to compare the MAS scores before and after the intervention in Experiments 1 and 2, with a significance level of 0.05. On observation of a significant interaction or main effect, the Wilcoxon signed-rank test was performed as a post-hoc test, adjusted using the Bonferroni correction for multiple comparisons. Statistical analyses were conducted using RStudio (version 2024.04.2+764) and SPSS (version 29.0; IBM, Armonk, NY, USA).

## Results

Statistical analysis revealed a significant main effect of time in Experiment 1 (F = 5.70, *p* < 0.05). Post hoc tests revealed that the MAS scores for VS and VS+AO significantly reduced after the intervention than that before, with no substantial difference between them (*p* < 0.05, Bonferroni-corrected), but not the AO score (Figure 3). Thus, VS significantly reduced MAS scores but not AO and AO did not enhance the effect of VS during VS+AO. In Experiment 2, significant main effects were observed for both intervention and time factors (F = 4.87, *p* < 0.05; F = 8.66, *p* < 0.01). Post hoc tests revealed that the MAS scores for VS and VS+MT significantly reduced after the intervention than that before, with no significant difference between them (*p* < 0.05, Bonferroni-corrected), but not the MT score (Figure 4). Thus, VS significantly reduced MAS scores but not MT, and MT did not enhance during VS+MT (Figure 4).

**Figure 3.**
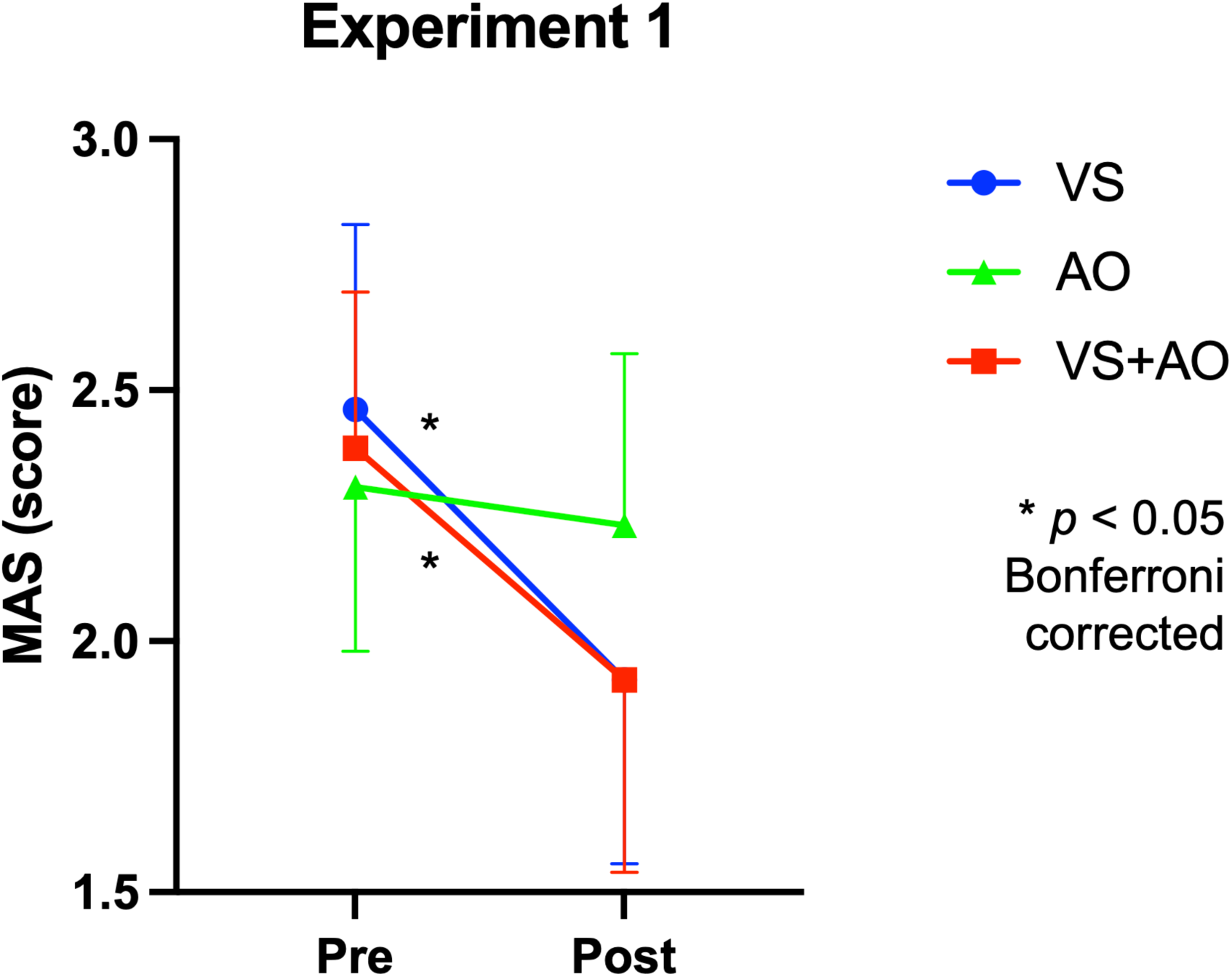
Comparison of MAS before and after VS, AO, VS+AO (Experiment 1). MAS, Modified Ashworth Scale; VS, Vibratory stimulation; AO, Action observation; VS + AO, Vibratory stimulation with action observation.

**Figure 4.**
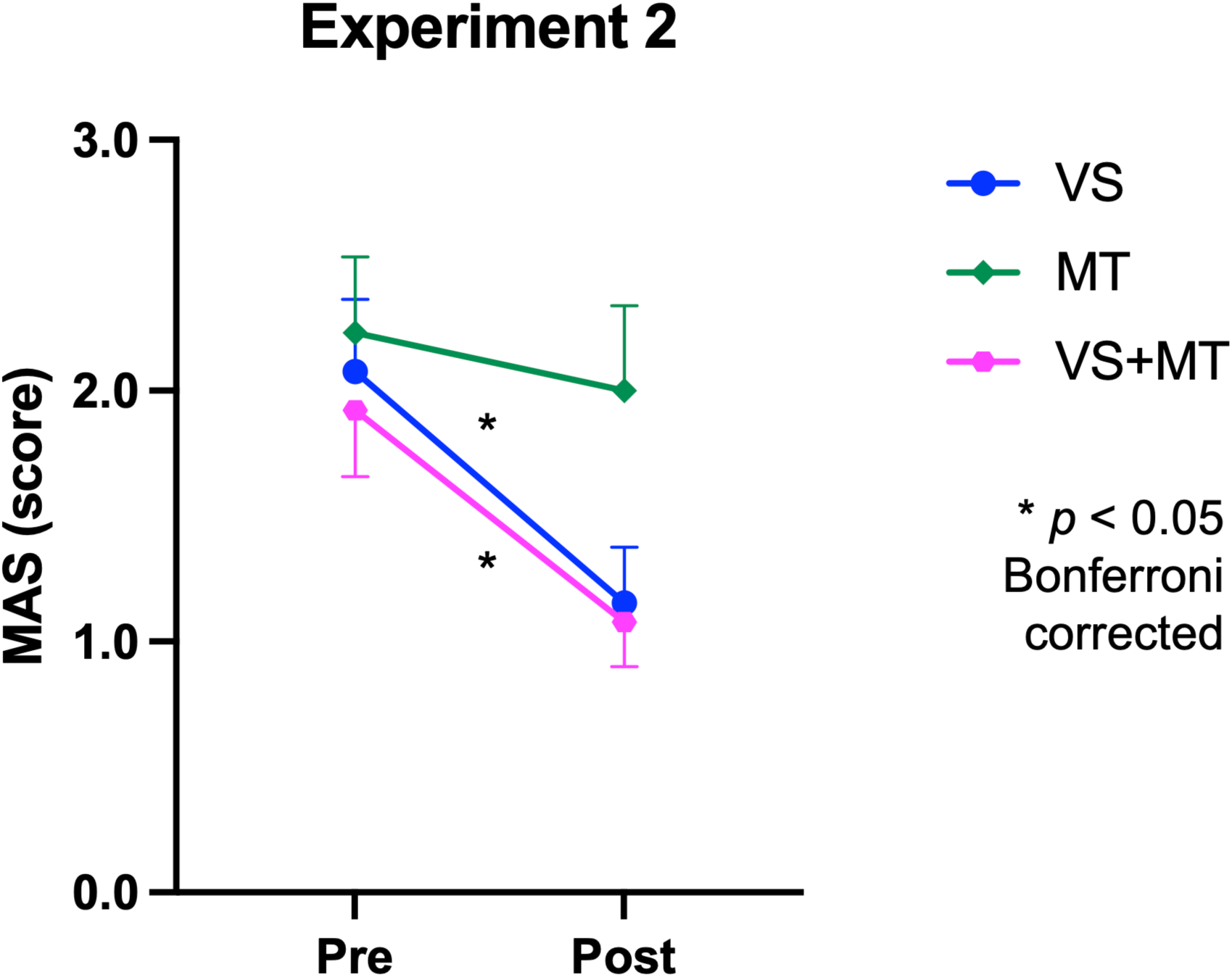
Comparison of MAS before and after VS, MT, VS+MT (Experiment 2). MAS, Modified Ashworth Scale; VS, Vibratory stimulation; MT, Mirror therapy; VS + MT, Vibratory stimulation with mirror therapy.

## Discussion

In this study, as hypothesized, 5-min of VS to the spastic muscles significantly reduced spasticity; while, AO and MT had no significant effect on spasticity. Furthermore, AO and MT did not enhance the VS effect during combined therapy (VS+AO or V+MT). Moreover, the present study included patients with chronic spasticity symptoms for many years. VS had an immediate effect, even in these patients, strongly suggesting its efficacy in reducing spasticity during the chronic phase.

### Why VS reduced spasticity but not AO or MT

Hand VS activates the muscle spindles, and the Ia afferent signals from the muscle receptors are sent to the brain (26–28). Our series of fMRI studies on hand VS have consistently revealed that this input activates the hand section of the primary motor cortex (M1) contralateral to the hand (29–32). However, M1 hand section activity is not reported during hand AO (33,34). Recent transcranial magnetic stimulation (TMS) studies have reported that AO alone does not always increase corticospinal excitability and when it does, the top-down motor imagery component is involved (25,35). As for MT, previous studies have reported significant activity in the precentral gyrus (premotor cortex or M1), ipsilateral to the moving hand (36) or not (37) during MT. Even in the former, the activity seems to be mainly located in the precentral gyrus anterior part, that is, most likely the premotor cortex, rather than the M1 hand section (36). Furthermore, motor imagery has been proposed to be actively combined for MT effectiveness in restoring motor function (38). Thus, VS can effectively reduce spasticity owing to its potential to directly affect the M1 hand region contralateral to the vibrated hand. The combination of VS with AO or MT did not enhance the VS spasticity reduction as neither AO nor MT substantially activates the M1 hand region.

### Possible mechanisms underlying spasticity reduction effect of VS

The mechanisms of spasticity reduction by VS remain unclear and require further research. Nonetheless, previous studies provide some insight into the mechanisms. Kimura et al. reported that, VS applied to the palms of mildly spastic hands, resulted in pronounced muscle activity in the extensor muscles than that in the flexor muscles of the fingers (39). As extensor muscle activity alleviates spasticity in the flexor direction, this activity may be involved in spasticity reduction. Embedded in the primate M1 is a long-loop reflex circuit via the cortex that receives Ia afferent signals from the flexor muscles and directs movement to the extensor muscles (40). The recruitment of this transcortical circuitry may be responsible for the appearance of remarkable extensor muscle activity on palm VS. More pronounced extensor muscle activity may have an inhibitory effect on the spinal cells innervating the flexor muscles through a reciprocal inhibition circuit at the spinal cord level. This does not contradict previous studies reporting that spastic muscle VS of the upper limb reduces the hyperexcitability of the spinal anterior horn cells (10,11).

Recent structural MRI studies have revealed that lesions in the superior and posterior corona, internal capsule, thalamus, putamen, precentral cortices, and insula may be associated with contralateral upper limb spasticity (41). These damages in the unilateral (contralateral) motor-related areas and motor descending pathways may cause interhemispheric imbalance, i.e. ipsilateral dominance, and this ipsilateral dominance of descending pathways from the precentral cortices ipsilateral to the paretic limb is proposed to be one of the causes of spasticity (42). Such spasticity may be mitigated by VS reducing the ipsilateral involvement, through interhemispheric inhibition from the contralateral precentral hand section activated during VS to the ipsilateral one (32).

Finally, it is shown that the basal ganglia (the putamen) is most frequently lesioned in patients with spasticity compared to those without (43,44). As the basal ganglia play a critical role in muscle tone control (45), this lesion may largely affect muscle tone. As hand VS activates M1 hand region and subcortical motor networks including the basal ganglia (putamen) and cerebellum (31), the M1-centered cortical and subcortical motor network interventions through VS may be involved in spasticity reduction. Owing to the higher sensitivity of the Ia afferent fibers with a thicker diameter to electrical stimulation, electrical stimulation of the hand muscles just below the motor threshold using a mesh glove may also reduce hand muscle hypertonia (46). Electrical stimulation of the hand muscles just below the motor threshold alters the functional connectivity between the putamen and the M1 (47,48). Thus, VS that likely stimulates the muscle afferents (see above) may also change the functional connectivity in the putamen-M1 circuit to reduce spasticity. Nevertheless, elucidating the exact mechanisms of spasticity alleviation by hand VS is necessary.

The study has some limitations. First, the study has a small sample size and did not include patients with different types of functional impairment or spasticity. Second, the VS stimulation site was limited to the fingers and palm, and only the flexion and extension movements of the fingers were used for AO and MT. Therefore, further research with large sample size is needed to assess similar effects of VS for body parts, targeting patients with different types of stroke.

## Conclusions

This study compared the immediate effects of VS, AO, and MT on spasticity reduction in the paretic hand of patients with chronic stroke and revealed that VS led to significant spasticity reduction, while AO and MT had no significant effect. Furthermore, AO or MT did not enhance the effect of VS in combined therapy. The results clearly demonstrate the effectiveness of VS rather than AO and MT in reducing the spasticity of the paretic hand. We propose the predominant clinical application of VS in reducing hand spasticity. However, we do not completely deny the possibility of spasticity reduction effect of AO or MT in some patients.

## Acknowledgments

We would like to thank the patients who participated in this study.

## Funding

This study was supported by JSPS KAKENHI Grant Nos. JP19H05723, JP23H03706, and JP23K17453 for EN, and by JSPS KAKENHI Grant No. JP23K10417 for HN, and by JSPS KAKENHI Grant No. JP23K19907 for KN.

## Declaration of interest statement

The authors report that there are no competing interests to declare.

## Data availability statement

Data supporting the findings of this study are available upon request from the corresponding authors. The data are not publicly available because they contain information that can compromise the privacy of the research participants.

## Notes

### Competing Interest Statement

The authors have declared no competing interest.

### Clinical Trial

UMIN000056192

### Author Declarations

This study was approved by the Kyoto Tachibana University Research Ethics Committee (approval number 21-47).

